# Impact of Vaccination and Testing in an Urban Campus model for the SARS-CoV-2 Pandemic ^*^

**DOI:** 10.1101/2021.02.02.21251040

**Authors:** Yi Zhang, Sanjiv Kapoor

**Affiliations:** Illinois Institute of Technology, Chicago, USA

## Abstract

The crisis induced by the Coronavirus pandemic has severely impacted educational institutes. Even with vaccination efforts underway, it is not clear that sufficient confidence will be achieved for schools to reopen soon.

This paper considers the impact of vaccination rates and testing rates to reduce infections and hospitalizations and evaluates strategies that will allow educational institute in urban settings to reopen. These strategies are also applicable to businesses and would help plan reopening in order to help the economy.

Our analysis is based on a graph model where nodes represent population groups and edges represent population exchanges due to commuting populations. The commuting population is associated with edges and is associated with one of the end nodes of the edge during part of the time period and with the other node during the remainder of the time period. The progression of the disease at each node is determined via compartment models, that include vaccination rates and testing to place infected people in quarantine along with consideration of asymptomatic and symptomatic populations. Applying this to a university population in Chicago with a substantial commuter population, chosen to be 80% as an illustration, provides an analysis which specifies benefits of testing and vaccination strategies over a time period of 150 days.

## 1 Introduction

The health crisis induced by the SARS-CoV-2 virus has resulted in severe disruption of college education, especially at residential universities. The shared space in residential colleges is a particular challenge and a study [14] reports on the beneficial impact of testing in this environment. While preventive measures that include lockdown have been used extensively [2, 12, 4], the measures have had substantial economic disruption and may not be effective in collegiate and school settings.

As the spread of the vaccine progresses, residential and urban colleges face even more economic challenges [13]. Even with vaccination in progress, the rate of vaccinations and the threat of new variants poses risks and challenges and a careful watch over the spread of the infection is critical. Even if the campus community observes safety protocols, ensuring safety is especially challenging for campuses in urban settings where there are commuter students as well as interactions with the city population. Testing often is a strategy that could work depending on the specificity and sensitivity of the tests [11, 17]. While testing students within campus will significantly ameliorate the spread of infections [14] most urban campuses have significant interaction with the surrounding area. Early detection of infections via testing in this scenario is even more important so that measures to protect the population in contact can be applied [15]. These considerations are even more important when considering the opening of K1-12 schools.

We consider an urban campus model that incorporates the impact of interactions with the urban population. While network models that incorporate the impact of travel between distinct locations have been considered [5, 6], these models do not account for a constant interactions of a fixed population, e.g. commuter students in a school, with a differentiated population, i.e. the city population. A distinction between the two sets of people is in the NPI strategies adopted, that include testing and physical distancing. Our interacting population model utilizes compartment models for interacting subsets of populations with each of the compartmental models based on earlier models developed in [16], which incorporate dynamic social distancing that modifies transmission rates as the fear of disease grows. This model adapted from the traditional SIR model[3, 7] as originated from the research of Kermack and McKendrick [8, 9, 10]), is termed the SIR-SD model. We account for the fact that the transmission rate of the virus would be higher for closed classroom and dormitory settings. Similarly, for K1-12 schools interactions amongst students could result in increased transmission rates.

Our research provides results on (i) the growth of infections within a small close-knit university campus community as well as potential hospitalization rates based on the interactions of the campus community with the surrounding city based on commuter students (ii) the impact of relative testing rates within and outside the campus community and (iii) the impact of the rate of vaccination.

We also provide an economic analysis based on costs of tests and estimates of hospitalization costs to identify strategies that would enable the universities to budget economic costs while keeping infections limited. The economic cost premises are based on earlier research in [14].

## 2 Methods

Our primary focus being the interaction between urban surroundings and the campus community we considered a graph model with the nodes representing the communities and the interaction between them by edges. In this scenario we use a simple two node model with a predetermined community of commuters associated with the edge between the two nodes and that share space with the two node communities during one of two periods of time within 24 hours. Considering a division of the daily 24 hours into two time periods, termed day and night, the commuter community is assumed to share space with the college campus during the day and with the surrounding community during the night.

The progression of the disease is modeled via compartment models with parameters that are node dependent. We incorporate vaccination rates and testing rates into our compartment models. We also incorporate the impact of release from lockdown. The compartment model further utilizes an asymptomatic population and provides compartments for hospitalized and non-hospitalized infected populations.

For our study we used a university setting in Chicago with substantial commuter student population. Model parameters were obtained from an earlier study [16] that incorporated the dynamic transmission rate and the impact of removal of lockdown.

### 2.1 The Multi-Node Testing Compartment Model

Our compartment model is defined in Figure 1 where transition between population sets are defined. The description of the population categories is defined in the table in Figure 1. In addition to the regular epidemic model which is used for the city population, we add testing and vaccination to the model corresponding to the campus and colored red in the diagram. The parameter *τ* is the daily testing rate and *ν* the daily vaccinating rate. *ζ* is the efficacy of the vaccine. Changes over time in the population categories are modeled by the following differential equations: For each node *v* in the graph we have:

**Figure 1:**
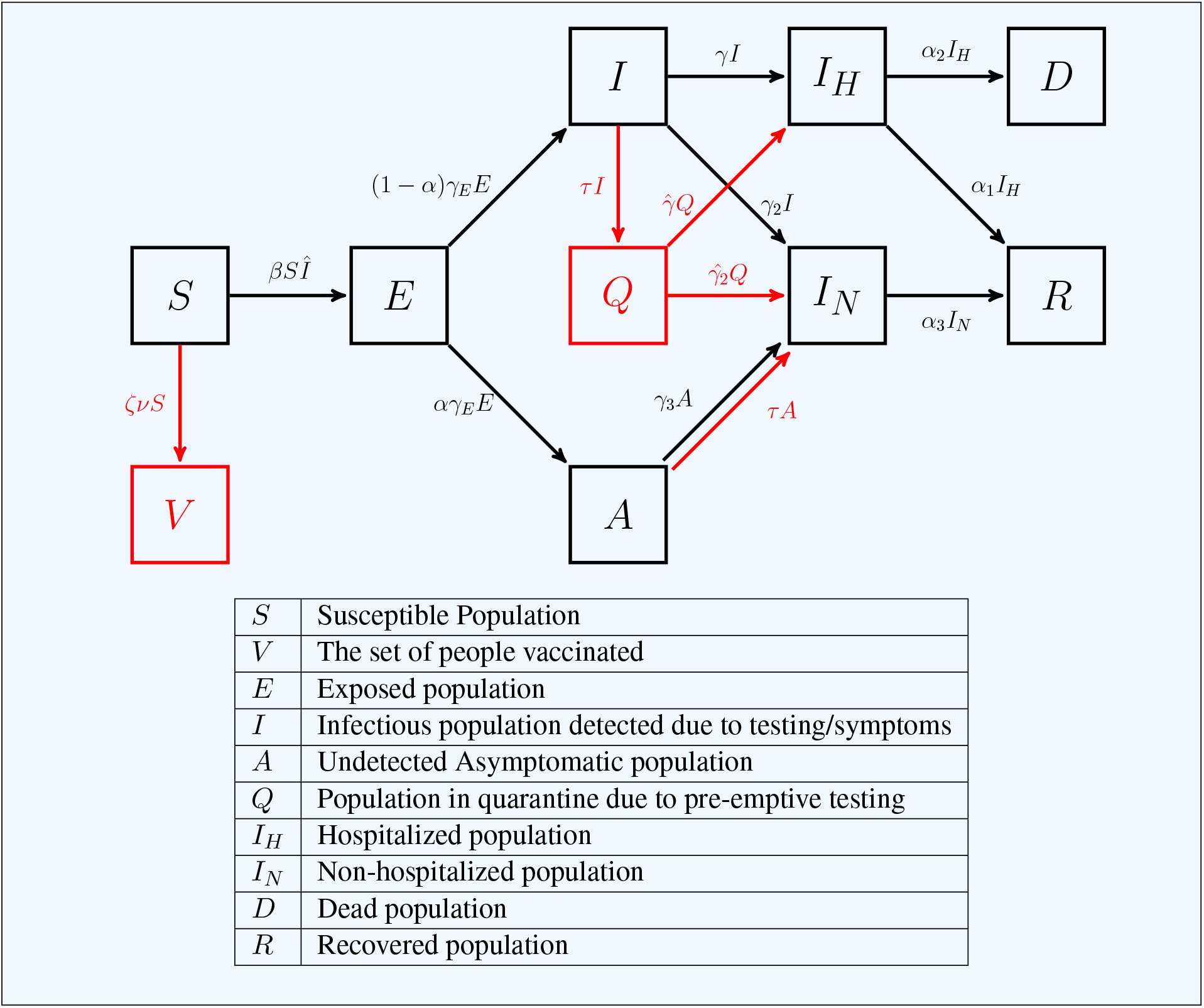
Transitions amongst population at node corresponding to the campus in the two-node SEAIR-SD Compartment Mode.

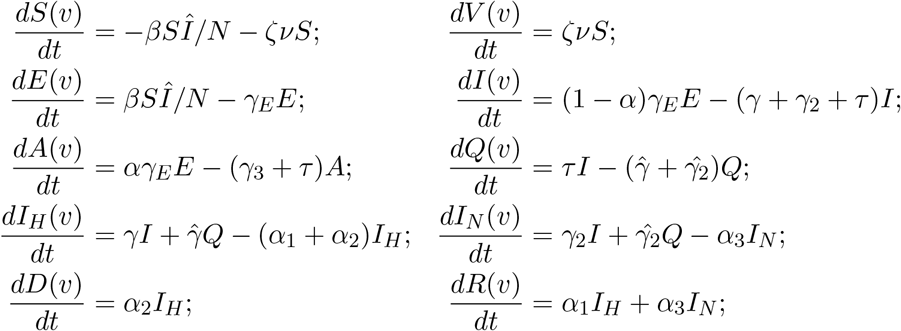

where *Î* = *I* + *A*.

In general transition between nodes will result in population transfer. In our case the population shift is based on the commuters and changes by the movement of the commuters every time period. In our simulation we assume 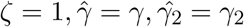. However, most likely these parameters would be altered in practise due to early intervention.

### Population Exchange

Consider a population exchange between the nodes *u* and *υ* over a 24 hour period via a fixed set of commuters. In this scenario we will consider two time periods, represented by *D* (day) and *N* (night), during which an exchange of population takes place between *u* and *υ*. Let us index the compartments *S, E, I* and *A* at node *u* as *S*(*u*), *E*(*u*), *I*(*u*), *A*(*u*). A similar indexing holds for the compartment populations at node υ. We let *S*_*C*_(*u, υ*) be the susceptible population that transfers between *u* and υ with corresponding terms *E*_*C*_(*u, υ*), *I*_*C*_(*u*, υ) and *A*_*C*_(*u*, υ) for the exposed, infectious and asymptomatic infectious commuter population. We will assume that the set of commuters is fixed. In our scenario commuter students travel from an urban campus (Node *u*) to the surrounding city (Node *υ*) in the evening (*N*) and return during the day (*D*). Each of the compartments are governed by the differential equations above and the dynamics of the disease are simulated during the day and night accounting for the changing population. The commuter population is added to the node *u* during *D* and to the population of node *v* during the period *N*. This results in the following governing equations for infections during *D*:

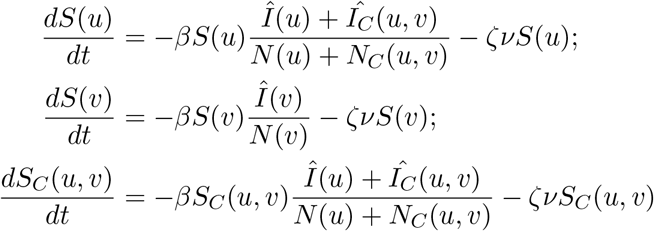

where *N* (*u*), *N* (υ) and *N*_*C*_(*u*, υ) is the total population at the nodes *u*, υ and the edge (*u*, υ), respectively. (Note that *Î* (*u*) = *I*(*u*) + *A*(*u*). Similar infectious population exists at node u and for the commuters).

The equation governing the dynamics during the night period *N* are:

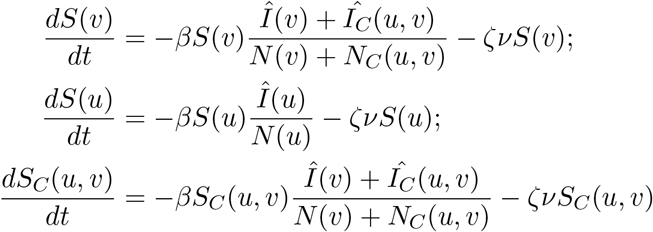

Other population dynamics are similarly defined.

## 3 Results

### Model Set-up and Parameters

Model specific parameters for the compartment model that we have defined are estimated by least square error fitting methods on a base model. These methods have been specified in [16] and the base model is contained in the appendix.

As a representative example, we utilized data from Cook county in Illinois, that includes Chicago, to obtain the parameters for our model. The model utilizes a time-varying transmission rate for the city, where a variety of measures can be adopted to reduce the transmission rate when the infection rate rises. Details of this dynamic transmission rate are provided in the appendix. For the node corresponding to the urban school campus we consider a fixed transmission rate (a contrast with dynamic social distancing parameters is planned), as it may be difficult to ensure all possible social distancing methods given the proximity in dormitories and classrooms. We assumed a representative susceptible population of 10,000 for the campus (note that in our model the entire population is not assumed to be susceptible), as compared to the population of Cook county which is 5,150,233. Our assumption in this study is that the commuter population is 80% of the school’s population. The testing rate of the county is set to 2%, which is an overestimate of average test performed to date [1], and that is kept constant in our model.

### Infection Rates and Hospitalization Rates

We consider the changes in infection rates and hospitalization rates for two distinct time partitions of 24 hours. For the first case we consider a time partition Π_1_, *D/N* = 1 where the time spent at the two nodes are equally partitioned and in the other case, time partition Π_2_, *D/N* = 1*/*2, where two-thirds of the time is spent in the city and one-third in the school. Different test rates were considered ranging from no test to 100% per day in steps of 20%. A testing rate of 20% corresponds to testing the entire school population once in 5 days.

The results show a marked decrease in infections and hospitalization cases. With a 20% testing schedule the number of infections drops to 40% of the original number of infections. The hospitalization numbers also show improvement though not in the same order of magnitude. The impact of early diagnosis has not been incorporated. This impact is illustrated for both Π_1_ and Π_2_ in Figure 2.

**Figure 2:**
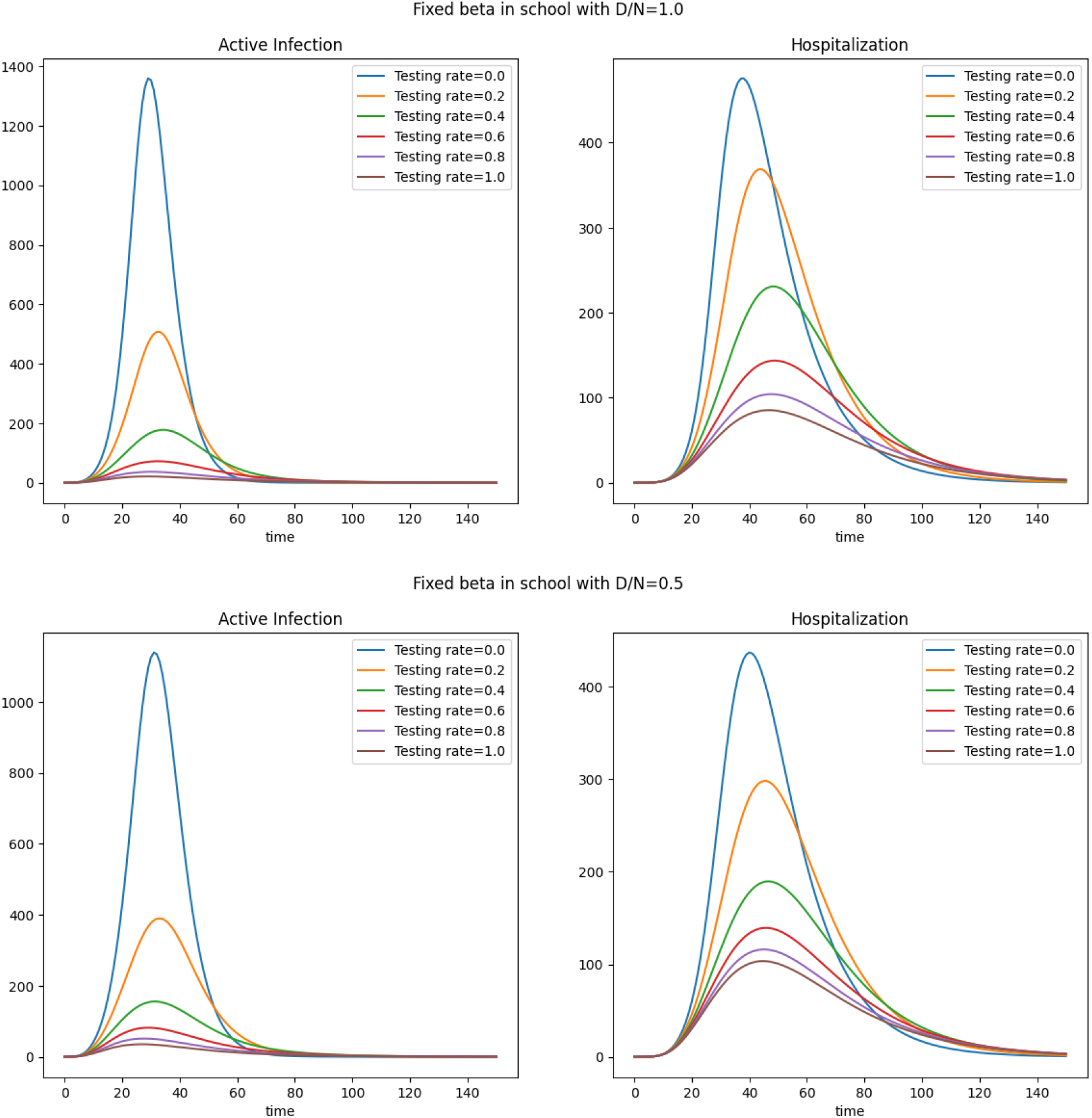
Peak infection and hospitalization under different testing rates with 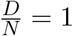 and with 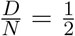.

### Combined Impact of Vaccinations and Testing

We also considered increasing the rates of vaccination and the rates of testing. As illustrated in the compartment model, vaccination reduces the size of the susceptible set and testing allows for faster isolation of infected cases thus reducing the spread of infection. The combined impact is illustrated in Figure 3, where at a testing rate of 14% (i.e. entire population tested roughly once a week) and vaccination rate of 0.4% of the population per day, the peak daily active infection rates (size of *I*) are reduced by 50% of the original numbers and peak daily active hospitalizations reduced by 20%. This does not account for the impact of early testing on hospitalization rates.

**Figure 3:**
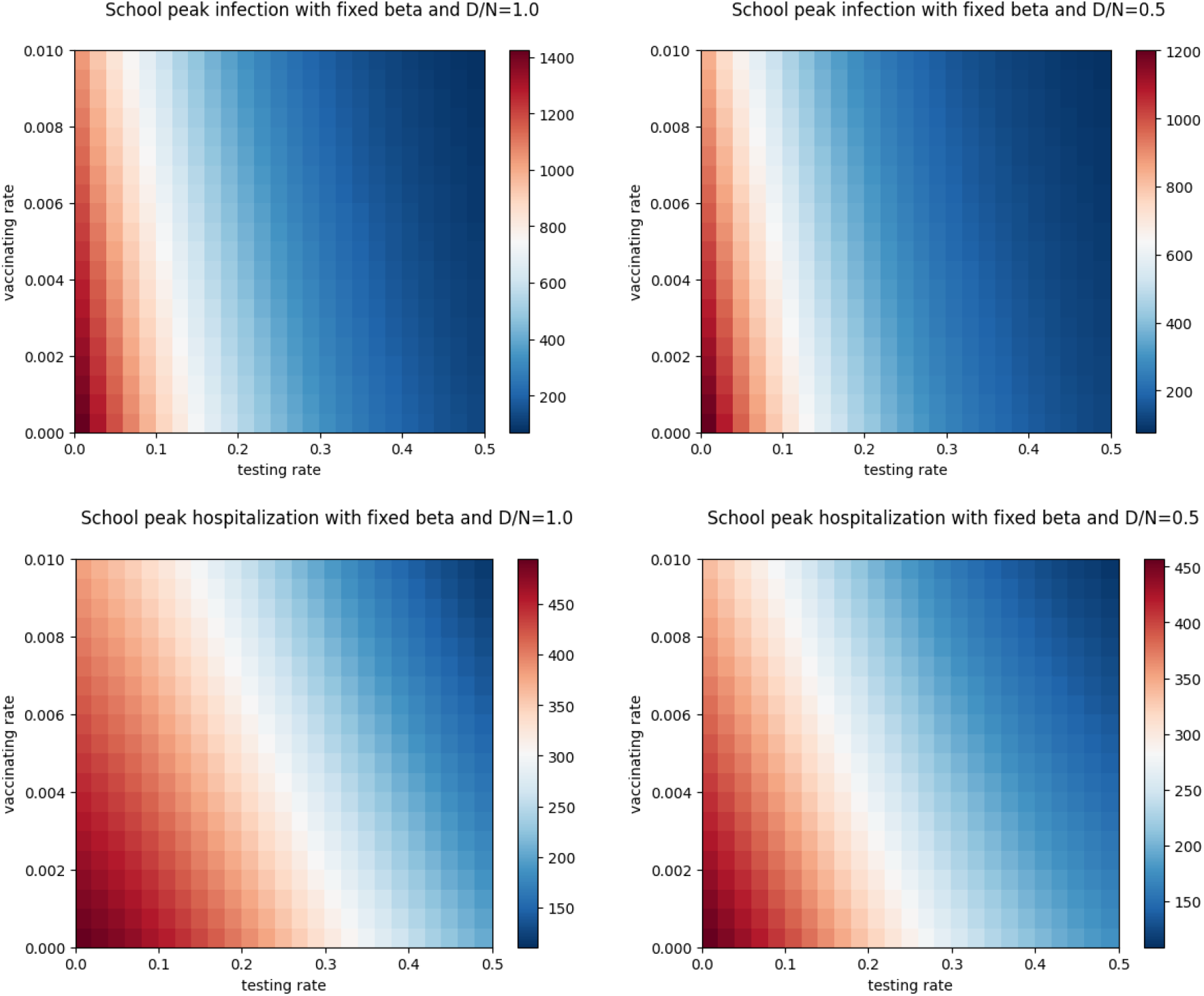
Grid of peak infection and hospitalization under different testing and vaccinating rates with 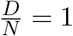 and 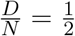.

### Cost Analysis

We also considered a cost benefit analysis of testing that results in reduced costs due to infections. Following the assumptions presented in [14] we consider the benefits per infection averted to be $5500, $8500, $11600 in the worst to best case. In the case of $11600 saving per infection, Figure 4 illustrates the total cost benefits in millions of dollars, peaking at a testing rate of 82.5% of population/day. Even a testing rate of once in 3 days will result in 10.5 million in savings for a school of population size 10,000, assuming that the daily vaccination rate is at 0.15% of the population. With a vaccination rate of 0.3%, doubled from previous, the improvements are even more, the saving increasing to 14.5 Million. Savings based on various test rates can be found in Table 1. The testing rates at which the saving becomes positive and the maximum savings can be found in Table 2.

**Table 1:** Savings for a range of testing rates under two vaccination rate scenarios. The savings are in millions of dollars for a population of size 10,000. All testing and vaccination rates are over 24Hr period.

|  | 0.15% vaccination rate |  |  | 0.3% vaccination rate |  |  |
| --- | --- | --- | --- | --- | --- | --- |
| Saving (\$/infection averted) | 5500 | 8500 | 11600 | 5500 | 8500 | 11600 |
| Every 2 weeks | -0.34559 | -0.20161 | -0.05284 | -0.19895 | 0.070072 | 0.348058 |
| Every week | -0.0782 | 0.598182 | 1.297107 | 0.363397 | 1.386965 | 2.444652 |
| Every 5 days | 0.552061 | 1.982283 | 3.460179 | 1.315977 | 3.329064 | 5.409254 |
| Every 3 days | 2.754163 | 6.820993 | 11.02338 | 4.416541 | 9.711371 | 15.18269 |
| Every 2 days | 4.974325 | 12.89656 | 21.08286 | 7.350449 | 16.97331 | 26.91693 |
| Everyday | -0.74323 | 12.93451 | 27.06818 | 1.230322 | 15.80622 | 30.86799 |

**Table 2:**
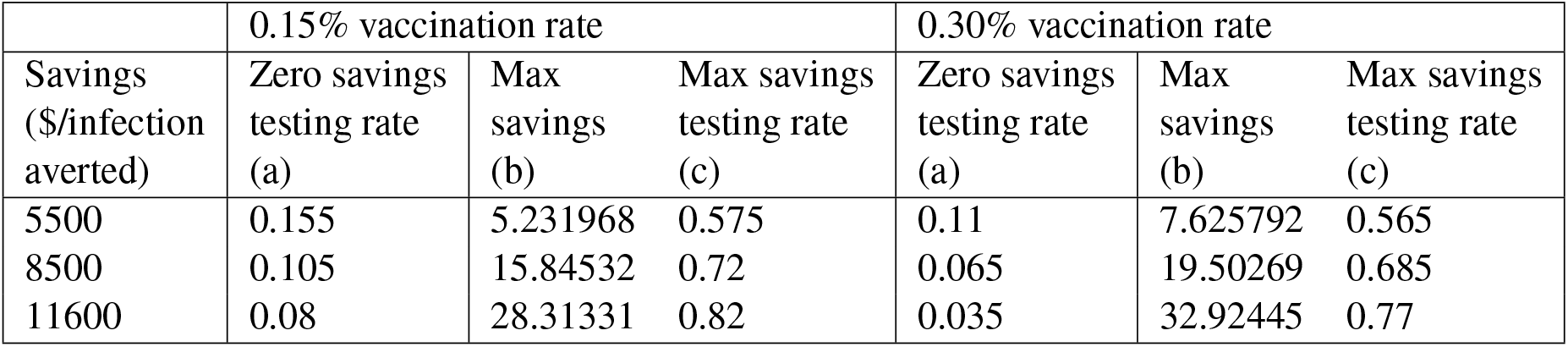
Table illustrating combination of testing rate that results in zero savings (no savings) in column (a) and maximum savings (column (b)) and corresponding testing rate (column (c)) for two different vaccination rates. Three Savings/infection averted dollar amount scenarios are considered. All savings are in millions and testing and vaccination rates are over 24Hr period. Savings are for a population of size 10,000.

|  | 0.15% vaccination rate |  |  | 0.30% vaccination rate |  |  |
| --- | --- | --- | --- | --- | --- | --- |
| Savings (\$/infection averted) | Zero savings testing rate (a) | Max savings (b) | Max savings testing rate (c) | Zero savings testing rate (a) | Max savings (b) | Max savings testing rate (c) |
| 5500 | 0.155 | 5.231968 | 0.575 | 0.11 | 7.625792 | 0.565 |
| 8500 | 0.105 | 15.84532 | 0.72 | 0.065 | 19.50269 | 0.685 |
| 11600 | 0.08 | 28.31331 | 0.82 | 0.035 | 32.92445 | 0.77 |

**Figure 4:**
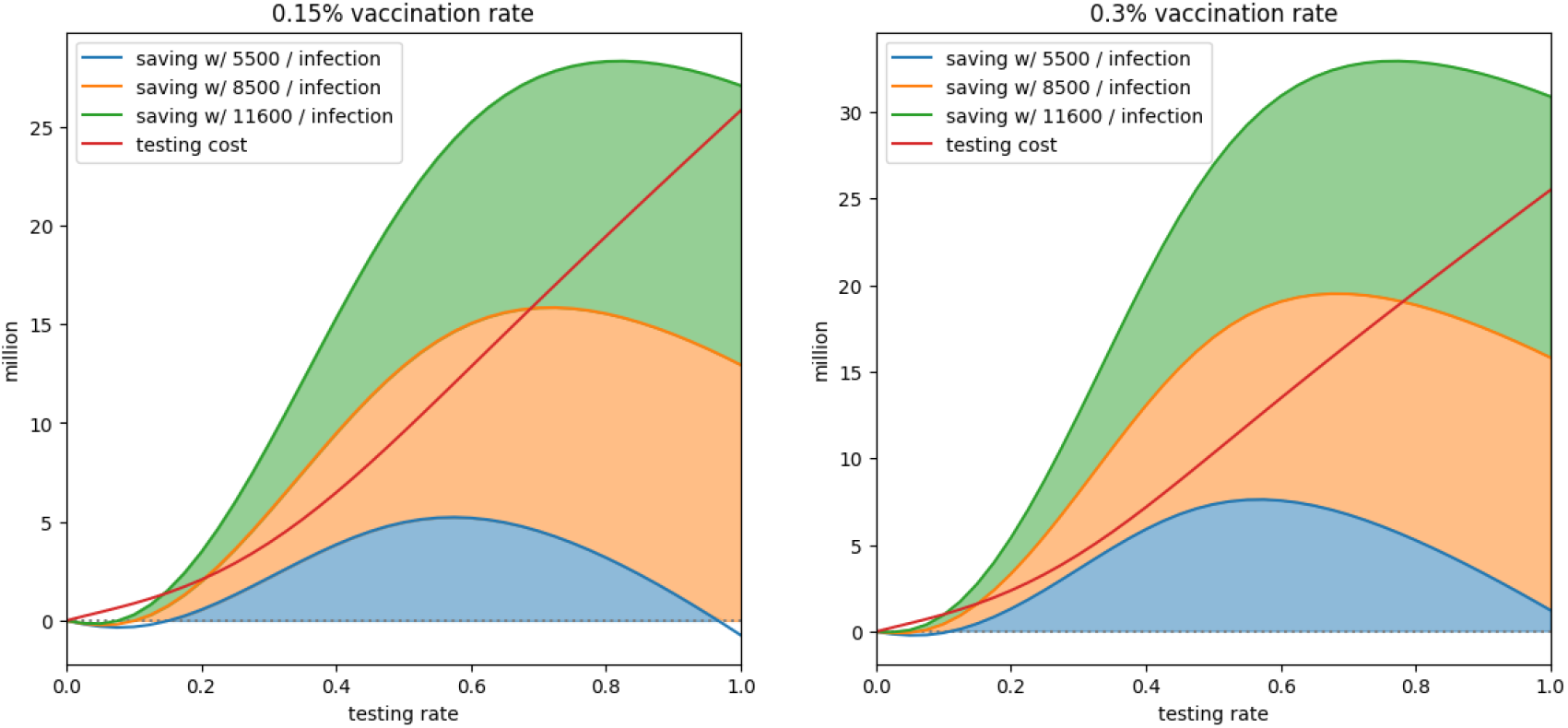
Total savings under 0.15% and 0.3% vaccination rate.

## Discussion

From the results it is evident that testing is very critical for re-opening schools even while vaccinations are ongoing. While increased vaccination rates would help it is unlikely that substantial fraction of the population will be vaccinated in less than 300 days. Our results indicate that for small population sets interacting with an outside population, it is beneficial to test frequently. A frequent testing schedule will result in overall economic benefits with savings increasing substantially as the tests get more frequent. Given the physical limitation, testing once in 2-3 days appear to be the most feasible and will result in substantial benefits.

## Data Availability

Data from public sources was used and referenced.
https://github.com/CSSEGISandData/COVID-19.

## 5 Acknowledgements

We acknowledge the use of cloud computing resources provided by the Chameleon project at Chameleon-cloud.org.

## Contributions

Both authors contribute equally. S.K designed the concept, the model and wrote the document, Y.Z participated in the model development, designed and wrote the code for the experiment and participated in the writing of the paper.

## 6 Supplementary Material

### Graph Model

The model we used is a special case of the network *G* = (*V, E*) shown in figure 5 where the nodes *C*_1_, *C*_2_ … ∈ *V* are community nodes while *S*_1_, *S*_2_ … ∈ *V* represent congregate places, e.g. schools. There are population sets at the nodes and also population set on edges that reside periodically, for a fixed fraction of the time period, on one or the other node *u* or *υ* corresponding to the edge *e* = (*u, υ*).

**Figure 5:**
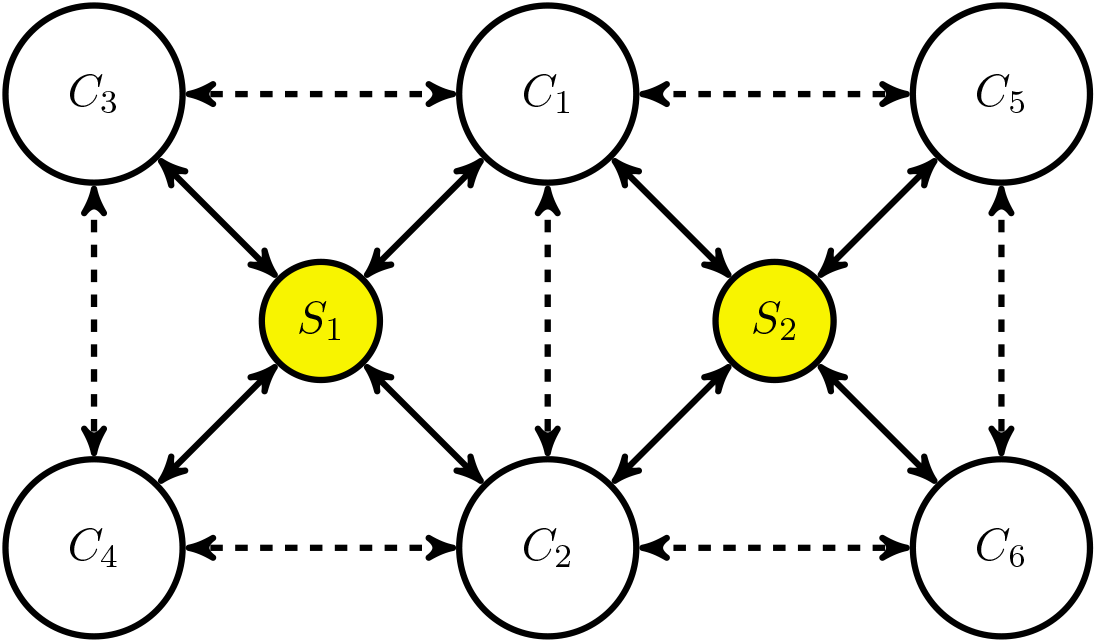
General graph model.

### Model Fitting for Cook County, Chicago

For determining parameters, we used a model description that is a variation of the model in [16] which did not include the tested and vaccinated populations.

The compartment model is presented in the Figure 6 where *H* represents the population that was under lockdown during the initial phases and that released the lockdown population into *S* over time.

**Figure 6:**
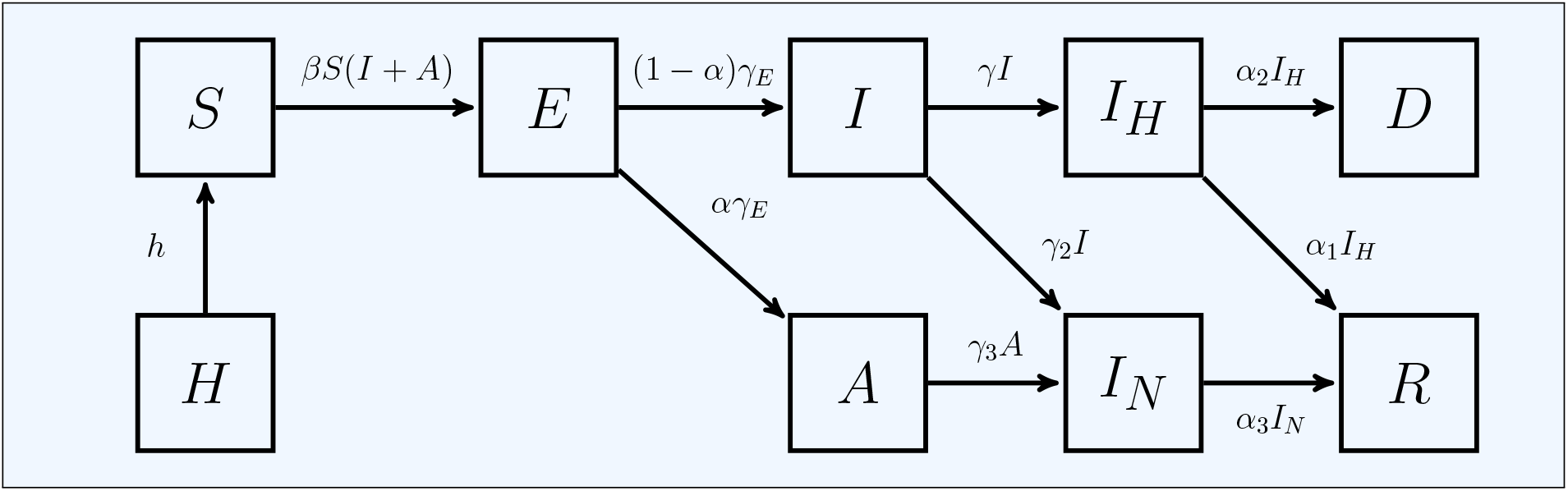
Transitions for a single node SIR-SD Compartment Model.

Using this model we estimated the parameters using data for Cook county in Illinois that contains the Chicago metro area. The fitting and the model parameters are shown in Figure 7. The transmission parameter *β* is dynamic and is defined in [16], stated here for easy reference:

**Figure 7:**
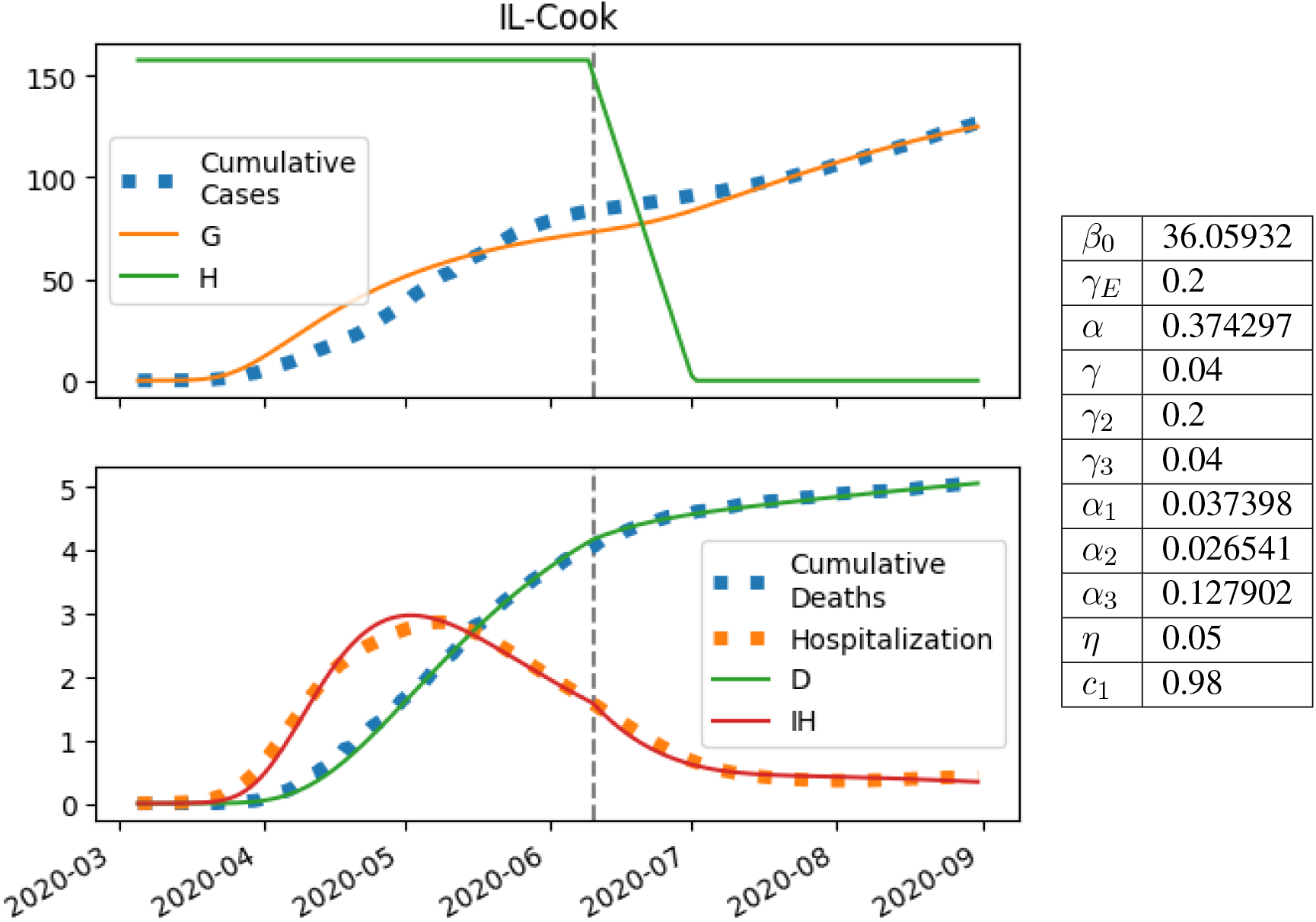
Fitting and parameters.

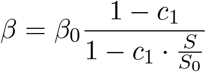

where *S*_0_ = *ηN* is the initial susceptible population and *β*_0_ a constant.

## References and Notes

[1] IDPH COVID-19 Statistics. Available at https://www.dph.illinois.gov/covid19/covid19-statistics.

[2] National Governors Association. Memorandum: reopening institutions of higher education. Available at http://www.nga.org/wp-content/uploads/2020/05/State-Higher-Ed-reopening-final.pdf.

[3] David W Berger, Kyle F Herkenhoff, and Simon Mongey. An SEIR Infectious Disease Model with Testing and Conditional Quarantine. Working Paper 26901, National Bureau of Economic Research, March 2020.

[4] Cornelia Betsch, Lars Korn, Philipp Sprengholz, Lisa Felgendreff, Sarah Eitze, Philipp Schmid, and Robert Böhm. Social and behavioral consequences of mask policies during the covid-19 pandemic. Proceedings of the National Academy of Sciences, 117(36):21851–21853, 2020.

[5] Serina Chang, Emma Pierson, Pang Wei Koh, Jaline Gerardin, Beth Redbird, David Grusky, and Jure Leskovec. Mobility network models of covid-19 explain inequities and inform reopening. Nature, 589(7840):82–87, 2021.

[6] Stephen Eubank, Hasan Guclu, V. S. Anil Kumar, Madhav V. Marathe, Aravind Srinivasan, Zoltán Toroczkai, and Nan Wang. Modelling disease outbreaks in realistic urban social networks. Nature, 429(6988):180–184, 2004.

[7] Herbert W Hethcote. The basic epidemiology models: models, expressions for R0, parameter estimation, and applications. In Mathematical understanding of infectious disease dynamics, pages 1–61. World Scientific, 2009.

[8] William Ogilvy Kermack and Anderson G McKendrick. A contribution to the mathematical theory of epidemics. Proceedings of the Royal Society of London. Series A, Containing papers of a mathematical and physical character, 115(772):700–721, 1927.

[9] William Ogilvy Kermack and Anderson G McKendrick. “contributions to the mathematical theory of epidemics. ii.—the problem of endemicity”. Proceedings of the Royal Society of London. Series A, Containing papers of a mathematical and physical character, 138(834):55–83, 1932.

[10] William Ogilvy Kermack and Anderson G McKendrick. Contributions to the mathematical theory of epidemics. III.—Further studies of the problem of endemicity. Proceedings of the Royal Society of London. Series A, Containing Papers of a Mathematical and Physical Character, 141(843):94–122, 1933.

[11] Joshua A Lieberman, Gregory Pepper, Samia N Naccache, Meei-Li Huang, Keith R Jerome, and Alexander L Greninger. Comparison of commercially available and laboratory-developed assays for in vitro detection of sars-cov-2 in clinical laboratories. Journal of clinical microbiology, 58(8), 2020.

[12] Polly Matzinger and Jeff Skinner. Strong impact of closing schools, closing bars and wearing masks during the covid-19 pandemic: results from a simple and revealing analysis. medRxiv, 2020.

[13] Kelly McLaughlin. The coronavirus could force smaller liberal arts and state colleges to close forever. Insider (April 2020). Available at https://www.insider.com/smaller-colleges-may-never-reopen-because-of-the-coronavirus-2020-4.

[14] A David Paltiel, Amy Zheng, and Rochelle P Walensky. Assessment of sars-cov-2 screening strategies to permit the safe reopening of college campuses in the united states. JAMA network open, 3(7):e2016818–e2016818, 2020.

[15] Rochelle P Walensky and Carlos Del Rio. From mitigation to containment of the covid-19 pandemic: putting the sars-cov-2 genie back in the bottle. Jama, 323(19):1889–1890, 2020.

[16] Yi Zhang and Sanjiv Kapoor. Hidden Parameters Impacting Resurgence of SARS-CoV-2 Pandemic. medRxiv, 2021.

[17] Wei Zhen, Ryhana Manji, Elizabeth Smith, and Gregory J Berry. Comparison of four molecular in vitro diagnostic assays for the detection of sars-cov-2 in nasopharyngeal specimens. Journal of clinical microbiology, 58(8), 2020.

